# Physical activity, sleep pattern and screen usage in undergraduate health-science students of Nepal – a cross-sectional study

**DOI:** 10.1101/2025.02.23.25322750

**Authors:** Jay Prakash Jha, Hiramani Prasad Chaudhary, Dilli Bahadur Pun, Lok Raj Joshi, Kapil Amgain

## Abstract

College students’ unhealthy habits of sleep and physical activity can adversely impact their overall well-being. Excessive internet usage has been linked to conditions like problematic internet use (PIU). However, there is less evidence on these issues and its correlates in Nepal. This study aims to investigate the patterns of computer usage, sleep habits, and physical activity among undergraduate health science students in Nepal, along with the frequency of PIU.

A cross-sectional study was conducted among Nepali undergraduate health-science students. Their sleep patterns (sleep score), physical activity levels (weekly MET value), computer usage habits, and problematic internet use (PIU score) were assessed through online questionnaire. Data were analyzed using descriptive and analytical statistics.

The study included 362 students (206 females), median age 21 years. A significant proportion of students exhibited inadequate sleep duration (30.45%) and experienced sleep disturbances frequently, such as early morning waking (23.48%) and racing thoughts in bed (23.38%). Most students reported minimal engagement in heavy exercise, with more than half (55.8%) classified as minimally active. Females were significantly less active than males (Chi Square 57.6, p<0.001). Screen usage were prevalent among students in smartphone and computer. PIU was observed in 56.94% students; the score was significantly associated with poor sleep quality (Spearman Rho 0.27, p<0.001), MET value (Rho −-0.237, p<0.001), daily duration of device and internet use (p<0.001). Regression analysis revealed sex (p=0.009), sleep score (p<0.001), MET value (p=0.001) and knowledge of computer technology (p=0.014) as significant predictors of PIU (R^2^=0.14).

This shows that poor sleep, low physical activity, and PIU are common among the students, and are closely interrelated. These issues can have consequences for academic performance, mental health, and overall well-being, making it essential to develop interventions to encourage healthy habits among undergraduate health-science students in Nepal.

**Author summary:** This study addresses a demanding issue of the relation between technology use and well-being of students. By inspecting the patterns of physical activity, sleep and internet usage among undergraduate health-science students, which provides the original insights that are both timely and significant. We have shown that the majority of students’ physical activity is low, have significant sleep problems and have high digital device usage, leading to high prevalence of problematic internet use.

This result highlight the significance of encouraging a healthier lifestyle choices in the era of digital academic fields. Our main goal is to inspire readers to adopt healthy lifestyle and be aware of excessive screen use. It also encourages other researchers to explore further on the causal relation between different lifestyle parameters and screen use. Practically, by slightly adjusting the screen time and exercise, it promotes the wellbeing of students and also their academic success. Overall, this study emphasise on role of health and education, driving the research in the area of public health.

## 1. INTRODUCTION

College students are at their significant stage of life where they are likely to have poor health practices. Sedentary behaviour, encompassing activities such as internet browsing, phone communication, and social media interaction, has multiple detrimental health effects such as metabolic syndrome, cardiovascular diseases, cognitive decline, and all-cause-mortality [1]. The proliferation of digital devices and internet connectivity globally underscores the escalating reliance on digital technology. As of 2023, approximately 64.4% of the global population enjoys internet access, with smartphones serving as the primary conduit for 92.3% of users. Alarmingly, the daily internet usage averages to six and half hours, significantly exceeding the recommended limits of 2 hours per day by the National Institute of Health [2]. Nepal is also seeing a growing internet penetration rate of 51.6%. Social media engagement emerges as a dominant online activity among Nepalese population, who spend an average of over 2 hours daily, predominantly via mobile devices [3,4].

In addition, a significant portion of medical students live independently and in hostels, which may contribute to a higher propensity for internet dependence [5]. Research indicates that internet dependence, akin to substance abuse or compulsive eating, can be objectively evaluated and manifests in various physical and mental health complications [6].

Dependency on digital gadgets causes various physical and mental issues [7]. Excessive and unhealthy use of internet has been studied for its effect in different dimensions of health, impacting academic performance and future career prospects. The concept of Problematic internet use (PIU) is used to explain a risky, excessive or impulsive nature of internet use, resulting in adverse consequences in physical, emotional, social, and functional dimensions of health [8]. Studies underscore the correlation between periods of heightened academic demand, inadequate sleep, and diminished academic achievement among university students [9]. Given the enduring impact of college lifestyle habits, comprehensive evaluation and monitoring of daily health-related practices are imperative for the formulation of targeted guidelines tailored to the students. Despite the critical need, there remains a paucity of research in our population concerning this multifaceted issue.

### 1.1 Objective

The study aims to examine the patterns of computer gadget usage, sleep habits, and physical activity, and to explore the interrelationships between them among undergraduate health-science students of Nepal.

## 2. METHODS

### 2.1 Sample

This was a cross-sectional study on the undergraduate health-science students of different colleges of Nepal. Six medical colleges were purposively selected representing different provinces of the country. Target population of this study was undergraduate health-science students enrolled in MBBS, BDS, nursing and paramedical sciences. All the consenting undergraduate students of different streams of health-science who responded to the survey were included in the study. Calculated minimum sample (z^2^pq/d^2^) was 333 taking p (prevalence of problematic internet use) of 31.9%,[10] for 95% confidence interval and 5% margin of error. Accounting for 10% attrition rate, we aimed for data collection from 370 students and finally, we were able include 362 students.

### 2.2 Questionnaire

Semi-structured questionnaire was prepared by literature review, and included the different habits of students within past 15 days. It consisted of 4 parts: (a) general demography, sleep time and sleep disturbances (modified from the Pittsburgh Sleep Quality Index) [11]; (b) Physical activity as per International Physical Activity Questionnaire (IPAQ) [12]; (c) computer usage habits (device used, duration of use of the device and internet, common uses of the gadgets, knowledge of computer technology, and usefulness of internet in study); and (d) Problematic Internet Use (PIU) Questionnaire developed from former PIUQ [13]. For scoring purpose, the normal sleep habit parameters were classified as 0 for normal and 1 for abnormal as per NSF guideline [14] and another study by Hirshkowitz et al [15]. The sleep disturbance responses were also scored as 0 for never or rarely, 1 for occasional and 2 for frequently or always. This score was added along with the scores of sleep habits to get the total sleep score ranging from 0 to 17. The Problematic Internet Use Questionnaire (PIUQ) included 14 questions about the frequency of the problems related to excessive internet use. It had a 5-point Likert scale response from 1 (for never) to 5 (for always). The score ranged from 14 to 70; higher score indicating more problem. To diagnose PIU in our study, the score’s median value 42 was chosen as cutoff point; participant scoring 42 or above was labelled as PIU positive. The questionnaire was modified from Demetrovics (2008) [16] based on the context of Nepalese students. They were later reviewed by experts as well and refined accordingly.

The questionnaire were built in jotform for distribution, a browser-based form that could be used in any device and submitted online. Students were approached via social media or phone calls, and the link to the form was sent. The CHERRIES guideline was followed for online survey [17]. No personally identifiable data were collected, but multiple submission from single user was prevented by automatic IP address analysis. The platform also used javascript to alert respondents on incomplete submission. Subjects with more than 10% of incomplete columns were rejected. All the information, the procedures of the study and rights to the participants were explained to each in person, via online forms or in phone before recruiting them. Participants’ consent were collected by online form which was to be submitted separately at the time of filling the form. Data were collected from January to August 2023. Ethical clearance was taken from Ethical Review Board of Nepal Health Research Council (Reg no. 499/2022 P, date 5 January 2023) before starting the study.

### 2.3 Covariates

Quantitative variables included: age, annual family income (in NPR), time of sleep and wake up, sleep duration (hours) sleep latency (minutes of duration taken for falling asleep from the time of bed), sleep score, number of MET-minute per week, degree of physical activity, years of internet use, daily hours of screen and internet use and PIU score.

Qualitative variables were gender; sleep problems; different uses of gadgets; different social media platforms; knowledge, interest and usefulness of gadgets; and awareness of privacy in online world.

### 2.4 Statistical Analysis

Descriptive statistics were used to count the frequency, percentage, mean, standard deviation, median and interquartile range. As the data were distributed mostly non-paremetric, analytical study was done using Wilcoxon Rank-sum test, Chi Square test, Spearman correlation, and linear regression statistics. A *p* value of less than 0.05 were taken as statistically significant at 95% significant level.

## 3. RESULTS

Total 362 students participated in the study, of which 156 (43%) were males. Their mean age was 22 (±2.34 SD), ranging from 19 to 32 years. Students belonged to wide variety of location, from Morang in east to Bajhang in west. Participants’ self-reported substance abuse habit was low: only 10 reported as smoker, 54 were occasional drinkers, and only one was heavy drinker. Four participants did not respond to the substance abuse question. The average income of their family was NPR 8,44,169±17,51,658 per annum, out of 279 participants answered. Age and annual income was not significantly different among different gender (Table 1).

**Table 1.** Frequency of different parameters in the participants and their comparison in male and female.

| Parameters | Median Total (IQR) | Median (IQR) Male | Median (IQR) Female | P value (Wilcoxon Rank-sum test) | Effect Size |
| --- | --- | --- | --- | --- | --- |
| Age | 21 (20 – 23) | 22 (21 – 23) | 21 (20 – 23) | 0.398 | 0.00 |
| Annual Income (x 100,000 NPR) | 5 (2 – 10) | 5 (2 – 8) | 6 (1.5 – 12) | 0.186 | 0.01 |
| Sleep time | 12 am (11 pm – 12 am) | 12 am (11:30 pm – 12:30 am) | 11 pm (10 pm – 12 am) | **<0.001\*** | 0.04 |
| Wake up time | 7 (6:30 – | 7 (6:30 – 7:30) | 7 (6:30 – 7:30) | 0.118 | 0.01 |
| (am) | 7:30) |  |  |  |  |
| Sleep duration<br>(hours) | 7 (6 – 8) | 7 (6 – 7) | 7 (6.5 - 8) | **0.001\*** | 0.03 |
| Sleep latency<br>(minutes) | 15 (10 – 30) | 15 (10 – 30) | 15 (10 – 30) | 0.297 | 0.00 |
| Wake up<br>frequency | 0.5 (0 – 1) | 0 (0 – 1) | 1 (1 – 2) | **0.02\*** | 0.02 |
| Sleep Score | 4 (3 – 6) | 4 (3 – 6) | 4 (3 – 6) | 0.388 | 0.00 |
| MET-minute per<br>week | 942 (459-1572) | 1389.5<br>(901.125 –<br>2478.75) | 652 (284.25 -<br>1110.25) | **<0.001\*** | 0.20 |
| Internet use<br>(years) | 7 (5 – 8) | 7 (5 – 8) | 7 (5 – 8) | 0.800 | 2.092e-4 |
| Screen use<br>weekdays (hours) | 4 (3 – 5) | 4 (3 – 5) | 4 (3 – 5) | 0.067 | 0.01 |
| Screen use<br>weekend (hours) | 6 (5 – 8) | 6 (4.5 – 8) | 6 (5 – 8) | 0.473 | 0.00 |
| Internet use<br>weekdays (hours) | 3 (2 – 5) | 3 (2 – 4) | 4 (2 – 5) | **0.010\*** | 0.02 |
| Internet use<br>weekend (hours) | 5 (4 – 7) | 5 (4 – 7) | 6 (4 – 8) | **0.025\*** | 0.01 |
| PIU score | 43 (37 – 50) | 43 (35 – 50) | 43 (37 – 50) | 0.565 | 9.406e-4 |
| * Statistically significant at 95% significance |  |  |  |  |  |

### 3.1 Sleep

The median sleep time for the students was 12 midnight and wake up time was 7 am, making seven hours as their median sleep duration. Four students did not respond to the sleep habit questions. Out of 358 respondents, about a third (109) got sleep for less than 7 hours, and 4 had more than 9 hours, which are abnormal for adults as per National Sleep Foundation [15]. Only 245 (68.44%) students had adequate sleep duration of 7 to 9 hours. Nineteen students had prolonged sleep latency of more than 30 minutes (maximum latency 2 hours) (Table 2).

**Table 2.** Sleep habits and disturbances in the participants (according to habit in past 2 weeks)

| Regular sleep habit |  |  |  |
| --- | --- | --- | --- |
| Parameters (n=358) |  | frequency | Percent |
| Sleep duration (hours)<br>(Range 4 to 12 hours) | 7 to 9 (normal) | 245 | 68.44% |
|  | <7 (inadequate) | 109 | 30.45% |
|  | >9 (excessive) | 4 | 1.12% |
| Sleep latency (minute)<br>(Range 0 to 120 minutes) | <30 (normal) | 339 | 94.69% |
|  | >30 (prolonged) | 19 | 5.31% |
| Wake up frequency at night<br>(Range 0 to 7) | 0 to 1 (normal) | 292 | 81.56% |
|  | >1 (frequent) | 66 | 18.44% |
| Sleep Disturbance |  |  |  |

| Sleep disturbances (n=360) | Never or rarely | Occasionally | Frequently |
| --- | --- | --- | --- |
| Trouble falling asleep | 159 | 168 | 33 |
| Thoughts racing too much while in bed | 100 | 179 | 81 |
| Frequent waking during sleep | 266 | 79 | 15 |
| Difficulty returning to Sleep | 194 | 134 | 32 |
| Early morning waking | 128 | 147 | 85 |
| Snoring | 291 | 57 | 12 |
| Excessive daytime sleepiness | 111 | 189 | 60 |

**Table 3.** Physical activity level in male and female students.

|  | Inactive | Minimally active | HEPA active | Chi Square | P value |
| --- | --- | --- | --- | --- | --- |
| **Male** | 23 (14.74%) | 100 (64.1%) | 33 (21.15%) | 57.6 | **<0.001\*** |
| **Female** | 98 (47.57%) | 102 (49.51%) | 6 (2.91%) |  |  |
| **Total** | 121 (33.43%) | 202 (55.8%) | 39 (10.77%) |  |  |
| * Statistically significant |  |  |  |  |  |
| HEPA, Health Enhancing Physical Activity |  |  |  |  |  |

The sleep disturbance were assessed by 7 item questionnaire, each had response in 3 point Likert scale: never or rarely, occasionally, and frequently. Two students did not respond in sleep disturbance questions. The early morning waking was the most common sleep disturbance followed by thoughts racing too much in bed. Snoring was frequently experienced only by 12 students.

For scoring purpose, each of the normal sleep habit parameters were classified as 0 for normal and 1 for abnormal. Each sleep disturbance response was also scored as 0 for never or rarely, 1 for occasional and 2 for frequently or always. Total sleep score were calculated by adding sheep habit score and sleep disturbance score, which ranged from 0 to 17. In the 356 participants answered completely, the median sleep score in our sample was 4. Although boys’ sleep time was more delayed and had lower sleep duration than girls’ and girls woke up more frequently than boys during their sleep, there is no significant difference in total sleep score between them (p=0.388, table 1).

### 3.2 Physical Activity

Physical activity level was assessed by International Physical Activity Questionnaire (IPAQ) where the students reported the amount and weekly frequency of different physical activity. Following formula was used to calculate the weekly MET value as per the activity (According to IPAQ interpretation protocol [12]):

MET by heavy exercise: minutes of exercise per day x 8 x number days per week MET by moderate exercise: minutes of exercise per day x 4 x number days per week MET by walking: minutes of walking per day x 3.3 x number days per week

Total MET value for each participant were counted by adding up the MET values by different activities. All students responded to the physical activity question. Based on the self-reported form, most of them did not engage in heavy exercise. Total mean MET value was 1323.19±1385.32 MET-min/week and median was 942 (IQR 459-1572) MET-min/week. Comparison between gender shows that boys had significantly more MET value than girls (p<0.001, table 1).

Physical activity level was classified according to IPAQ protocol as inactive, minimally active and Health Enhancing Physical Activity (HEPA) level active. More than half of the participants were minimally active (202, 55.8%). Boys were significantly more active than girls (table 1 and 3).

### 3.3 Screen use

Out of the 357 participants answered, all used gadgets of some form. The most common gadgets students used were smartphone (329, 92.16%) and PC (desktop or laptop, 170, 48.18%). More than half students (187, 51.66%) possessed multiple devices. Only 2 did not use smartphone while 69 did not use any PC regularly. The most common primary use of smartphone was scrolling some social media platform, followed by watching videos and movies. PC was used most commonly for reading and social media (Figure 1).

**Fig 1.**
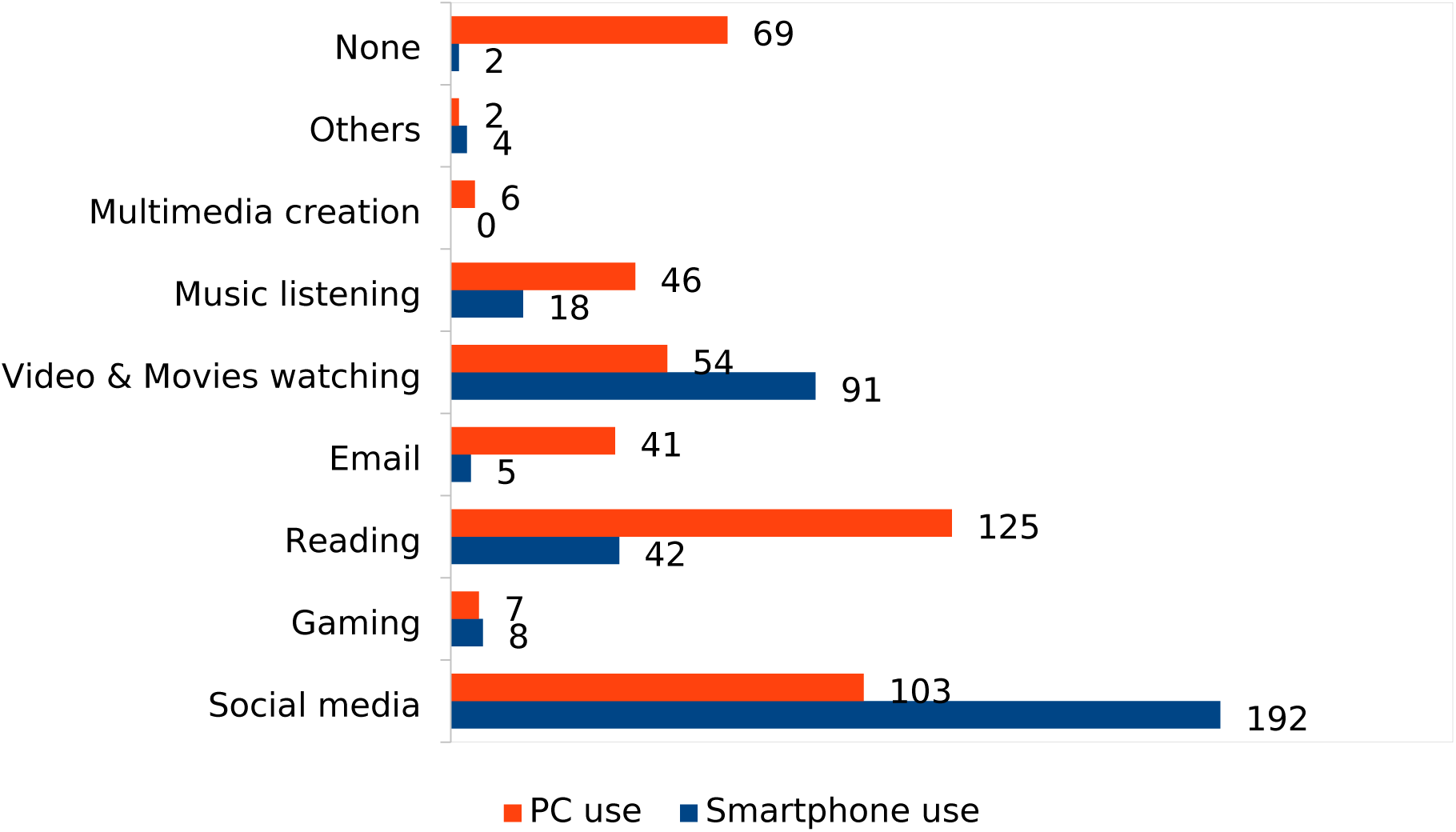
Common uses of gadgets in the participants (multiple response).

The most popular social media used by the students were Facebook/messenger and YouTube. Only one student reported not to use any social platform. (Figure 2)

**Fig 2.**
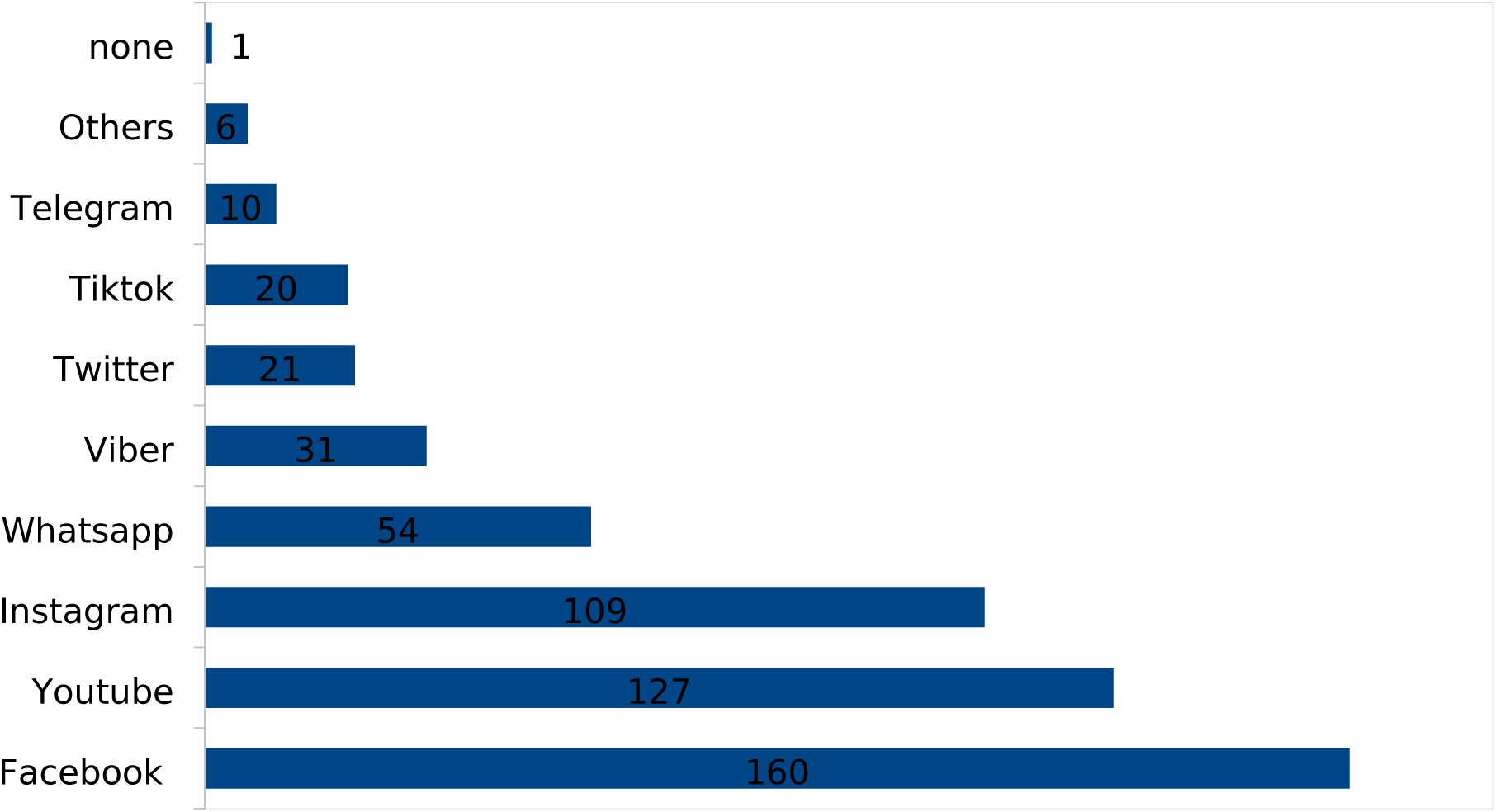
Frequency of common social media platforms used by participants (multiple response)

### 3.4 Daily device and internet use

The mean duration of any device (screen) use in the 357 students was 4.16±1.88 (maximum 12) hours per day on weekdays, which rose to 6.36±2.62 (maximum 16) hours per day on weekends and holidays. On an average, students were using internet over past 6.87±3.33 years. Their daily internet consumption rate was 3.79±2.03 (maximum 12) hours, which bumped to 5.79±2.75 (maximum 18) hours on weekends and holidays. Eighty percent (289) students regularly looked at screen for more than 2 hours daily. Girls used internet significantly longer every day than boys (p=0.01 for weekdays and 0.025 for weekend, Table 1).

### 3.5 Knowledge and utility of the devices

About a third (32.32%) students had poo (nil to beginner)level of knowledge in computer system, but 56.25% expressed their high interest in learning it. Only a quarter of students were aware of the value of their private data in internet and practised securing their personal data regularly. About 85% participants found the internet “very useful” or “were dependent on it” for study (table 4).

**Table 4.** Knowledge, interest and usefulness of computer technology.

| Computer use parameters | Frequency | Percent |
| --- | --- | --- |
| **Knowledge (n=362)** |  |  |
| Nil | 17 | 4.70% |
| Beginner | 100 | 27.62% |
| Working level | 155 | 42.82% |
| Above average | 73 | 20.17% |
| Very good | 17 | 4.70% |
| **Interest (n=352)** |  |  |
| Not at all | 3 | 0.85% |
| A little | 44 | 12.50% |
| Moderate | 107 | 30.40% |
| Much interested | 164 | 46.59% |
| Eager to learn | 34 | 9.66% |
| **Awareness of risk of privacy and practice of maintaining privacy in internet use (n=352)** |  |  |
| Not aware at all | 22 | 6.25% |
| Heard, but not cared | 13 | 3.69% |
| Heard, but not practised | 87 | 24.72% |
| Aware and practise sometimes | 142 | 40.34% |
| Well aware and practise regularly | 88 | 25.00% |
| **Usefulness of internet in study (n=352)** |  |  |
| Nil | 0 | 0.00% |
| A little | 11 | 3.13% |
| Moderate | 42 | 11.93% |
| Very much | 243 | 69.03% |
| Dependent on internet | 56 | 15.91% |

Table 5 describes the correlation between multiple pairs of parameters. It shows that amount of daily screen and internet use is positively correlated with higher sleep score (lower sleep quality). The amount of internet use in holidays is negatively correlated with total MET score over a week.

**Table 5.**
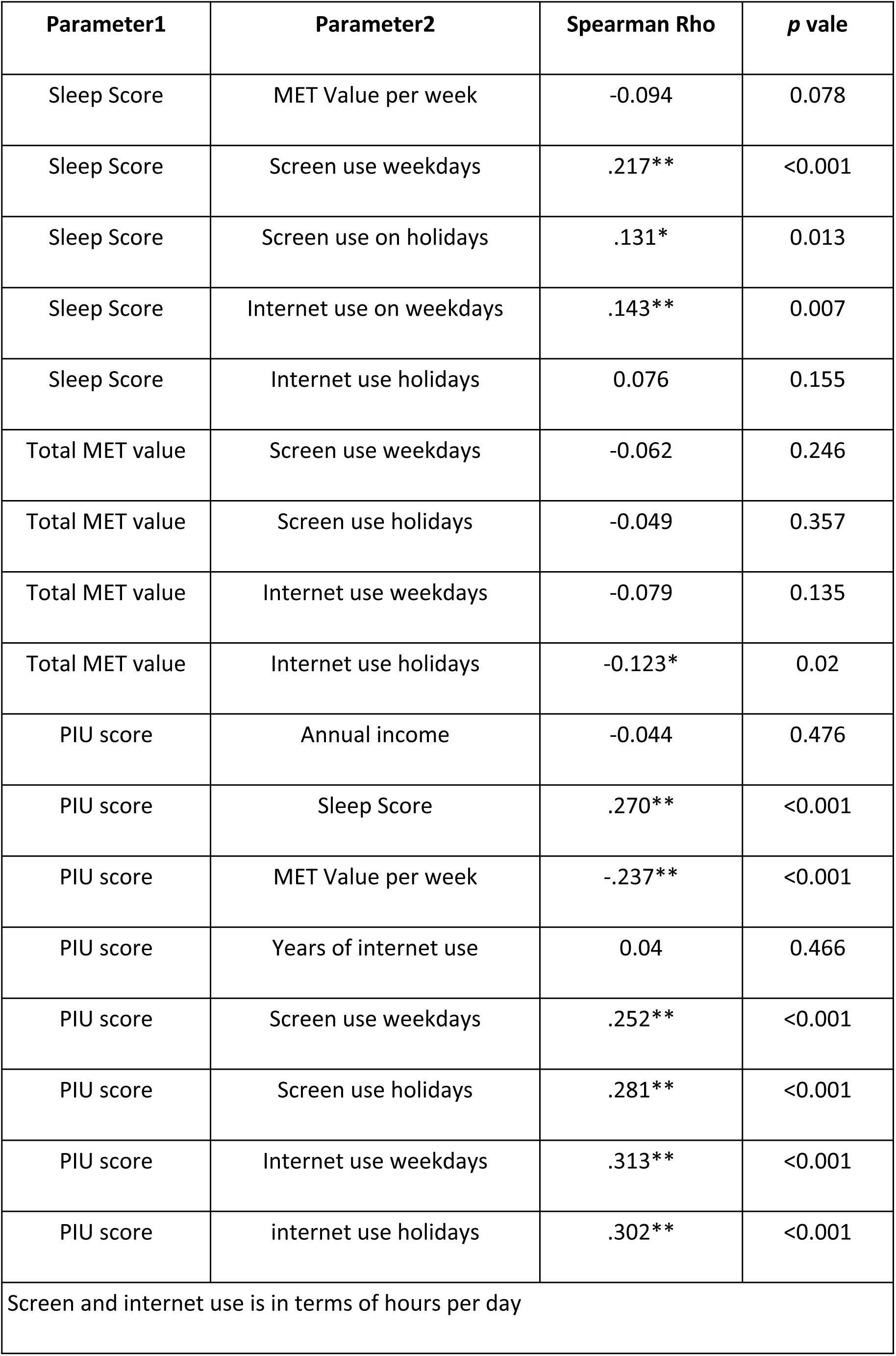
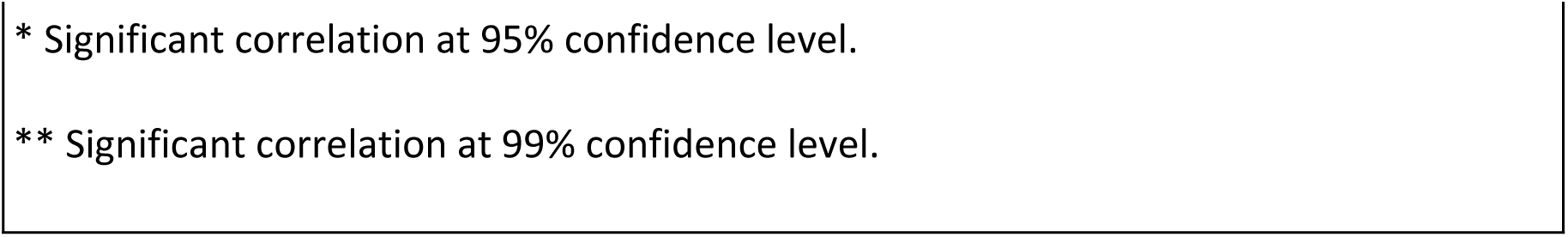
Correlation of different health parameters.

| Parameter1 | Parameter2 | Spearman Rho | p vale |
| --- | --- | --- | --- |
| Sleep Score | MET Value per week | -0.094 | 0.078 |
| Sleep Score | Screen use weekdays | .217** | <0.001 |
| Sleep Score | Screen use on holidays | .131* | 0.013 |
| Sleep Score | Internet use on weekdays | .143** | 0.007 |
| Sleep Score | Internet use holidays | 0.076 | 0.155 |
| Total MET value | Screen use weekdays | -0.062 | 0.246 |
| Total MET value | Screen use holidays | -0.049 | 0.357 |
| Total MET value | Internet use weekdays | -0.079 | 0.135 |
| Total MET value | Internet use holidays | -0.123* | 0.02 |
| PIU score | Annual income | -0.044 | 0.476 |
| PIU score | Sleep Score | .270** | <0.001 |
| PIU score | MET Value per week | -.237** | <0.001 |
| PIU score | Years of internet use | 0.04 | 0.466 |
| PIU score | Screen use weekdays | .252** | <0.001 |
| PIU score | Screen use holidays | .281** | <0.001 |
| PIU score | Internet use weekdays | .313** | <0.001 |
| PIU score | internet use holidays | .302** | <0.001 |
| Screen and internet use is in terms of hours per day |  |  |  |

\* Significant correlation at 95% confidence level.
\*\* Significant correlation at 99% confidence level.

### 3.6 Problematic Internet Use (PIU)

Problematic internet use questionnaire included fourteen question about their internet use, each with 5 point Likert scale; higher scale indicating more problematic use of internet. The score ranged from 14 to 70. The median score of the scale, 42, or higher was classified as PIU positive; and score less than 42 was classified as PIU negative.

Three hundred fifty three students responded the PIU questionnaire completely. The mean PIU score was 43.21±9.28, with median 43 (IQR 37-50), ranging from 18 to 70. More than half (201, 56.94%) got the PIU score 42 or higher. The PIU score did not differ significantly between boys and girls (p=0.565) (Table 1).

The PIU score was found to have mild but significant positive correlation with sleep score and daily screen and internet use. PIU score correlated negatively with MET value per week of activity. It was not related to annual income and years of internet use (Table 5).

Multiple linear regression analysis was conducted to explore the predictors of problematic internet use (PIU) among the students. The model included age, sex (coded as 0 for female and 1 for male), income, sleep score, total weekly MET value and knowledge on computer, as predictors (Table 6). It demonstrated a statistically significant relationship with the PIU score as dependent variable (F(6, 261) = 7.14, p < 0.001). The overall model explained 14% of the variance in PIU scores (R^2^ = 0.14, adjusted R^2^ = 0.12).

**Table 6.**
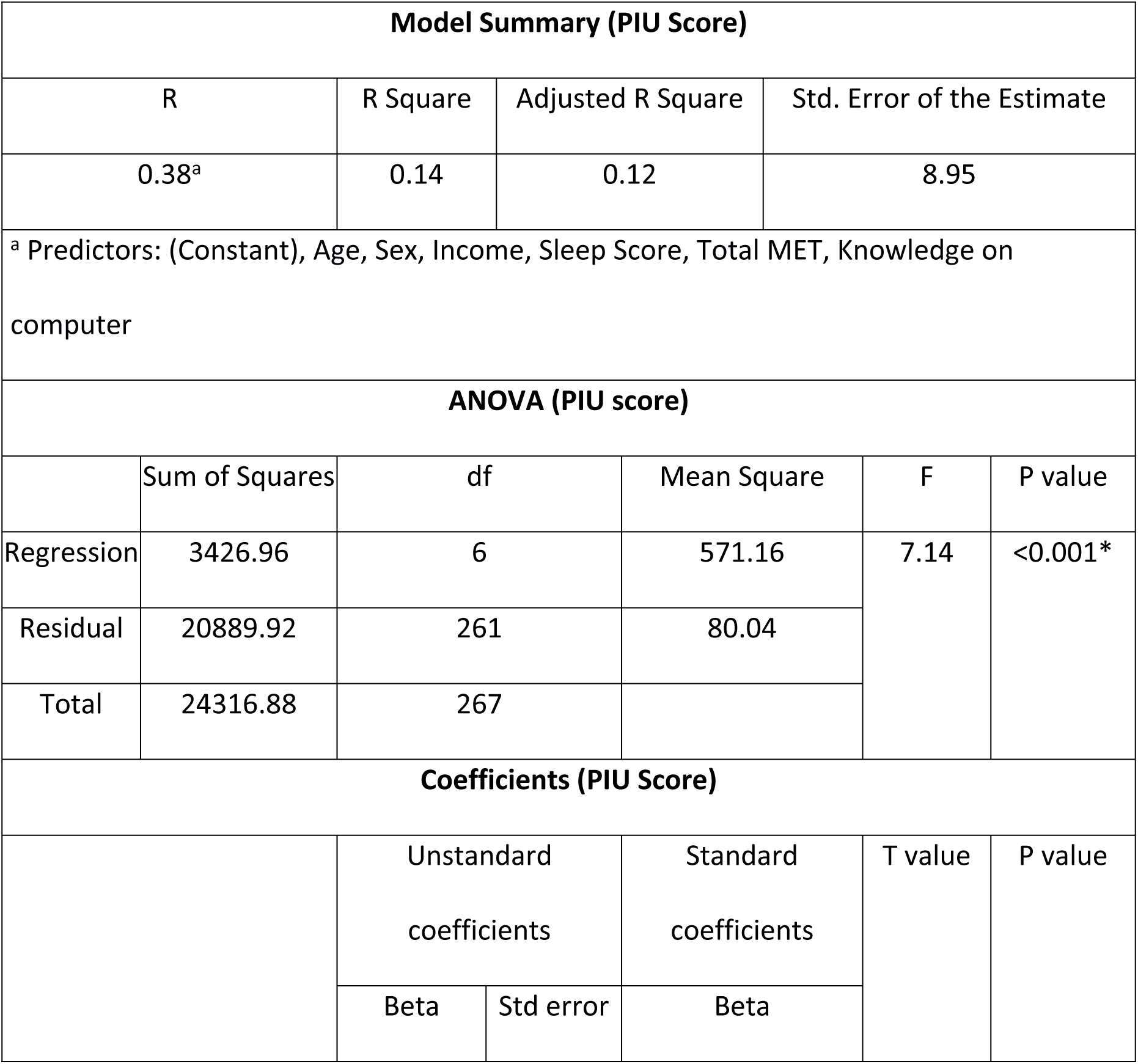
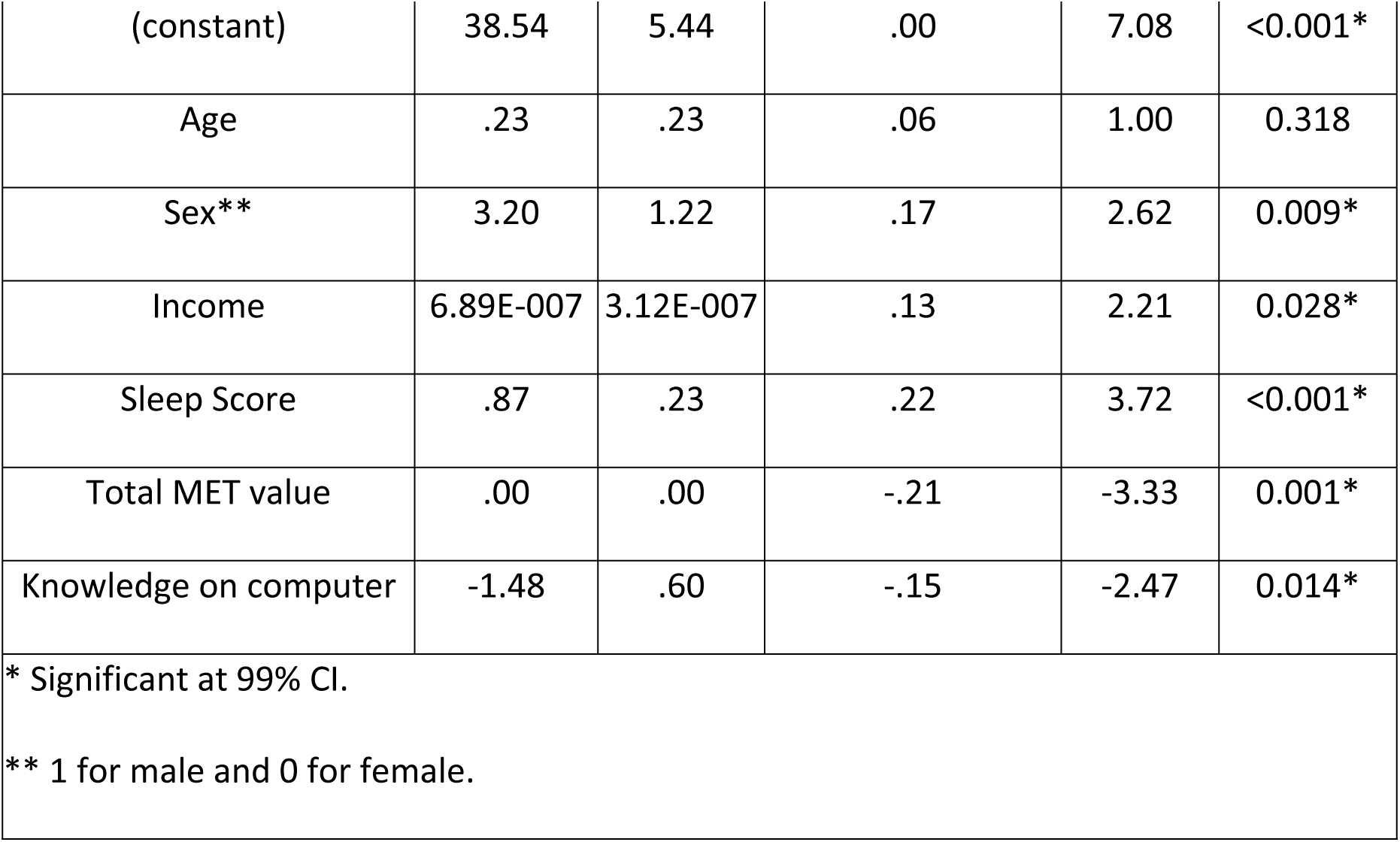
Regression analysis for predictors of PIU score.

Among the predictors, sex (β = 3.20, p = 0.009), sleep score (β = 0.87, p < 0.001), total MET (β = - 0.21, p = 0.001), and knowledge on computer (β = −1.48, p = 0.014) emerged as significant predictors of PIU. Male students tend to have a higher PIU score by 3.2 units compared to females (significant at p = 0.009). Similarly, higher sleep score was associated with increased PIU scores. In contrast, higher levels of physical activity and higher tech knowledge were associated with lower PIU scores. Higher income levels were also associated with higher PIU scores (β = 6.89E-007, p = 0.028), indicating that family income may have minimal effect in the score. These findings suggest that sex, socioeconomic status, sleep habits, physical activity levels and knowledge on computer play roles in problematic internet use among undergraduate health-science students.

## 4. DISCUSSION

This study shed light on the interrelationship between screen device usage habits, sleep quality, physical activity levels, and problematic internet use (PIU) among undergraduate health-science students of Nepal. Our analysis revealed several notable patterns and associations that have implications for understanding and addressing PIU in this population.

### 4.1 Sleep

Our study describes the sleep habits among undergraduate health-science students, revealing concerning trends in sleep duration and quality. While the median sleep duration was 7 hours, approximately one-third (30.45%) of participants reported inadequate sleep duration, consistent with findings in medical students of Kathmandu [18], which showed the figure to be 31.5%. However, our sample exhibited a lower proportion of poor sleep quality overall (10.4%) compared to other studies in Nepal (35.4%) [19]and in India (60%) [20]. This discrepancy may be attributed to differences in the study population and the questionnaire tools. Additionally, geographical variation or seasonal variation may be responsible for the differences [21–23]. For instance, it can be assumed that people living in urban area have access to different recreational facilities that keep people indulging outside whereas it will not be possible in rural areas. Also, people tend to go to bed early and wake up late in winter season compared to summer.

We found a significant differences in sleep timing and duration between boys and girls; which is similar to other literature explaining that women have earlier sleep time owing to their shorter circadian clock, and that they interpret sleep quality differently than men [24–26].

Various studies have described the importance of sleep quality and quantity in various domains of life, including academic performance in students [9,27]. Oliver et al (2018) [28]demonstrated that sleep quality and duration was the best predictor for quality of life and perceived physical wellness in US college students.

In our study, sleep quality was only one among the multiple health parameters related to computer use, and hence we did not use the full Pittsburgh sleep quality index. Future study could utilize it in a longitudinal study design, specially addressing sleep issues in wider population would yield a better picture about the gender difference in sleep parameters.

### 4.2 Physical activity

Our study revealed concerning trends in physical activity levels among undergraduate health-science students. Only 10.77% of participants engaged in health-enhancing exercises, and over half were classified as minimally active, aligning with recent findings indicating insufficient physical activity among university students [29]. This echoes global concerns, with WHO (2022)[30] reporting that over 80% of adolescents and 27% of adults worldwide fail to meet recommended physical activity levels.

In contrast, a study based on nationwide STEPS Survey data of 2013 in 4143 Nepali adults of 15 to 69 years age using Global Physical Activity Questionnaire has revealed that more than 96% of Nepalis have recommended levels of physical activity [31]. This high prevalence is not shared by other studies. For example used the GPAQ in 945 high school students to find that about 20% students had inadequate physical activity [32]. This discrepancy in prevalence of activity among the studies and our finding may be due to difference in sample population and the tools used. This might indicate a concerning trend of lower physical activity in young population, which might increase the proportion of non-communicable diseases in the future. Further investigation in different target groups of Nepali population might provide better picture in this section.

Sex differences in physical activity were evident in our sample, with males had higher activity levels compared to females, consistent with previous research [33–35]. However, conflicting findings exist, as studies like Bertrand *et al* observed higher physical activity among females during the COVID-19 pandemic [36]. Luo and Zhong observed the differential trend of changing behaviours in US students about physical activity and sedentary life while emerging from adolescent to adulthood [37]. Thus there seems to be influence of multiple contextual factors, cultural norms, societal pressure and individual preferences that contribute to the apparent discrepancies in the gender difference in physical activity in our study.

The relationship between sleep quality or duration and physical activity has been a subject of interest in various studies. Studies have shown that physical activity has positive effect on sleep quality [38–40] and both physical activity and sound sleep have positive effect on health-related quality of life [41]), and higher life satisfaction [42]. Our findings showed no significant correlation between these factors. Although the association between sleep and exercise have been discussed in multiple studies [43–48], the assumption that sleep quality or quantity can be enhanced solely through physical activity has been challenged [49–53]. This discrepancy is likely due to multiple confounders between these variables. Furthermore, studies typically take account of duration and time of sleep, not addressing its quality. It appears that physical activity and sedentary time have independent effect in sleep quality and/or quantity. Future study might utilize a separate investigation of sedentary time along with physical activity to comprehensively understand its effect in sleep quality as well as quantity.

### 4.3 Screen use

Our study provides insights into the patterns and implications of screen use among undergraduate health-science students. Participants reported average daily internet use of 3.8 hours in weekdays and 5.8 hours in holidays, with 80% spending two hours or more on screens daily. While our findings indicate lower screen time than the global average [3,4], it is higher than studies in Indian population [54,55] and in Jumla [56]. This highlights significant reliance on digital devices among college students, particularly for social interaction and information-seeking purposes.

The association between screen time and various health outcomes is well-documented in literature, as excessive screen use is linked to poor sleep, cardiovascular diseases, stress, and psychological problems [7,55,57–60]. While this association varies across different context [59], [61]), a meta-analysis showed strong and consistent association between use of devices and reduced sleep quantity and quality, as well as increased daytime sleepiness [62]. Consistent with this, our report has shown the daily screen and internet usage of the participants to be negatively correlated with sleep quality.

### 4.4 PIU

Our study highlights the prevalence and correlates of problematic internet use (PIU) among undergraduate health-science students, with 57% of participants exhibiting PIU. Prevalence of PIU in literature may differ with the population of study, time-frame and methodology, and ranges from 1.6 to 43.3% [19,56,58,63]. PIU is a measure of problems caused by misuse of internet, addressing the problems of social, behavioural, and emotional issues. It assesses the factors such as salience, excessive use, neglect of work, anticipation, lack of control, and neglect of social life. PIU is a broader term than internet addiction, which is characterised by loss of control and feeling of withdrawal [8,16]. Although we did use a cut-off value in the score for classification of PIU, it is essential to understand that that it is of less significance than the absolute score, as the goal is to minimize the score value.

A study by Balhara et al in 2019 included 2634 participants from 8 countries, among which Nepal had the highest prevalence of PIU (12.6%) [58]. The authors used Generalized Problematic Internet Use Scale-2 (GPIUS2) for assessing PIU, which is different from ours; but another study by Pawan Sharma et al [10], using the GPIUS2 scale on Nepali medical students done in 2018 found the prevalence to be 32%. Another Nepali study done in college students of Chitwan and Kathmandu found the prevalence of internet addiction to be 35%, and that it mediated the effects of sleep quality on depressive symptoms [19]). In line with literature findings, our study demonstrates PIU score to be associated with higher sleep score, higher daily device and internet use and lower physical activity [47,54].

Although the PIU score was not significantly different between males and females in our study, adjusting other factors shows being male is independently associated with a higher PIU score than females (β=3.20, p=0.009). While some literature shows PIU, internet addiction or dependency to be more common in males [54,58,64] including in Nepali population [10,19], others report females to have more PIU [18]). Such gender-mediated differences in internet addiction tendencies are due to economic factors, internet availability, social norms and some addiction-related health factors [65].

Usage of portable devices means that users are almost constantly connected and exposed to the risk of its excess use. The anxiety related to fear of missing out (FOMO) is the psychological manifestation of dependency on the devices. It is also causally linked to psychological issues such as suicidal tendency, depression, negativity and cognitive impairment [7]. A high screen time also increases stress [66,67], affects students’ academic performance [68,69], and health-related quality of life [70]. In a study on students of Cairo, the sleep quality and academic performance were correlated negatively with night-time screen use, and positively with physical exercise [71]. Another study in China demonstrated that the combination of high screen time and insufficient vigorous physical activity was associated with the high prevalence of psychological problems such as depression, anxiety and school life dissatisfaction[72].

Our study adds to this body of literature by highlighting the association of PIU with knowledge of computer technology. A significant proportion of our students reported inadequate computer knowledge, but are very interested in learning it. A lower knowledge in technology is significantly associated with higher PIU score (β=-1.48, p=0.014). Inadequate technical skills can impede students’ effective use of devices for studying, while excessive device usage may also have detrimental effects on their academic performance. Considering a high dependence on the internet reported by 85% of the participants for their studies, it is important to balance the adequate digital knowledge and appropriate device usage.

### 4.5 Recommendations

Recognizing the low physical activity level, poor sleep quality, high screen use and high PIU score, it is important to implement intervention to the young health-science students. The sleep quality needs to be improved by incorporating sleep hygiene education and fostering healthy sleep habits. The institutes also need to consider increasing physical activity opportunities on campus, integrate active breaks into academic schedules and offer fitness programs. Moreover, the negative correlation between computer-related knowledge and PIU implies that enhanced familiarity with technology may function as a protective factor against PIU. This study encourages the policymakers in training workshops on responsible internet use. This way, institutions can cultivate a culture of responsible internet use and empower students with essential skills for digital responsibility.

### 4.6 Limitations and future directions

As a cross-sectional design, this study precludes establishing causal relationships between variables. The reliance on self-report measures introduces the potential for recall bias and social desirability effects. Additionally, the study focused solely on undergraduate health-science students, limiting generalizability to other populations. Finally, while efforts were made to control for confounding variables, there may be unmeasured factors influencing the relationships between variables.

In future research, it is important to examine the patterns of internet usage and prevalence of problematic internet use (PIU) across different population groups. Additionally, longitudinal studies can provide insights into the effects of excessive internet use on sleep quality among undergraduate health-science students. Furthermore, the influence of socioeconomic factors on problematic internet use warrants exploration, along with strategies to mitigate its impact. Future research should also delve into the mechanism underlying the contribution of technology knowledge to PIU, offering valuable insights for intervention development and prevention efforts. Finally, intervention programs aimed at reducing PIU should be further investigated for their effectiveness.

## 5. CONCLUSION

A high degree of physical inactivity, computer gadget use and problematic internet use have been observed in the undergraduate health-science students of Nepal. As Nepal’s technological progress exposes its youth to a rapidly evolving digital environment, unregulated use of computer gadgets may lead to unhealthy practices. Promotion of Interventions for promotion of sleep hygiene and physical activity, minimization of screen usage while maintaining adequate digital literacy and responsible internet use need to be implemented at personal and institutional level. Future research with a prospective study design including diverse sample could provide further insights into the complex relationship between PIU and its predictors.

## Data Availability

Data will be available on request.

## Acknowledgement

Authors would like to acknowledge the important contribution of Mrs Bobby Thapa and the participants/undergraduate students from different institutes of Nepal.

